# Acute kidney injury and hyponatremia in hospitalized patients with rotavirus infection

**DOI:** 10.1101/2025.07.21.25331655

**Authors:** Ulrike Hoffmann, Antje Rückner, Olaf Nickel, Kathrin Marx, Ralph Wendt

## Abstract

**Introduction:** Rotavirus is a common cause of infectious gastroenteritis in infants and children. The role of rotavirus infections in adults has potentially been underappreciated and there is a paucity of data on incidence and outcome of acute kidney injury in adult patients.

**Methods:** We conducted a retrospective cohort study of adult hospitalized patients with microbiologically confirmed rotavirus infection. The primary outcome was occurrence of acute kidney injury related to rotavirus infection. Secondary outcomes were in-hospital mortality, duration of hospitalization and occurrence of sodium disorders.

**Results:** 314 hospitalized adult patients with rotavirus infection were evaluated. 200 patients, (63.7%) had community-acquired and 114 patients (36.3%) had nosocomial rotavirus infection. Acute kidney injury (AKI) occurred in 127 (40.4%) patients. AKI occurred more often in patients with community-acquired than nosocomial infection (110 (55.0%) vs 17 (14.9%), p<0.001). 26 (8.3%) patients died in hospital. Patients with AKI had worse survival (HR 2.63 (CI 1.20, 5.74) p=0.01). Hyponatremia was detected in 60 (30.6%) of 196 patients with community-acquired infection. Dehydration occurred in only 5 (2.6%) patients.

**Conclusion:** Adult outpatients with rotavirus infection and certain risk factors (age > 70 years and comorbidities, e.g. CKD) have a high risk of developing AKI. Patients should seek medical attention with a low threshold and, if necessary, undergo hospitalization to counteract volume depletion and the development of acute renal injury. Hyponatremia frequently occurs while dehydration is rare. Recommendations in outpatients at risk for AKI should focus on increasing salt intake rather than water intake.

## Introduction

As a major pathogen of viral gastroenteritis, rotaviruses repeatedly cause severe episodes of watery diarrhea, which is often accompanied by hospitalization, not least as a zoonotic and nosocomial infection. Infants under the age of five are particularly affected, but also immunosuppressed and immunosenescent patients. The role of rotavirus infections in adults has potentially been underappreciated. Rotavirus infection in adults typically manifests with nausea, malaise, abdominal pain, diarrhea, and fever[1]. While lethal cases are hardly recorded in Germany, up to 215,000 children still annually die in developing nations as a result of acute rotavirus infection[2]. The underlying pathomechanisms are not completely understood, but the expression of the viral enterotoxin NSP4 has been identified as one of the main reasons for the development of osmotic diarrhea[3].

The replication site and target cells of rotaviruses are the matured enterocytes of the villus tips, first of the duodenum, and later also of the jejunum and ileum. The extent to which rotaviruses cause systemic, extraintestinal infections was controversial for a long time. However, with the establishment of more sensitive molecular biological analysis methods, it was possible to detect both the viral particles and the nucleic acid in the liver, kidneys and the central nervous system, among others [4,5]. Although it is now considered certain that the host cell tropism of rotaviruses is extensive and that a large number of target tissues can be infected [6], possible rotavirus-associated end-organ damage is still too rarely considered in everyday clinical practice.

Despite the fact that hospitalized patients with rotavirus infections are in need of administration of electrolytes and the substitution of fluids in most cases, the influence on kidney function is insufficiently recorded. Only a few case reports in infants and children have shown that severe gastroenteritis can lead to impairment of renal function [7–9]. In addition, there are rare case reports of glomerulonephritis and acute kidney failure in children as a result of rotavirus infection [10,11].

Because of the paucity of data on incidence and outcome of acute kidney injury in adult patients with rotavirus infections, the aim of the present project is therefore the retrospective analysis of incidence of acute kidney injury (AKI) and renal outcome in adult patients with acutely detected rotavirus type A infection.

## Methods

### Study Design and Cohorts

This retrospective, single-center cohort study complied with the Declaration of Helsinki and was approved by the Ethics Committee of the German-Saxonian Board of Physicians, Dresden, Germany (number, EK-BR-86/23-1). Adult patients hospitalized at the St. Georg Hospital Leipzig between 2012 and 2023 with microbiologically confirmed rotavirus infection and at least two serum creatinine measurements were included in this study. Children and adolescents, outpatients and patients with detection of additional intestinal viruses (e.g. noroviruses, astroviruses and adenoviruses) were excluded from the study. Clinical information, laboratory parameters and course of disease were derived from the medical history of each patient. Comorbidities were extracted from the diagnosis-related group (DRG) coding data. Acute kidney injury (AKI) was graded using the KDIGO Acute Kidney Injury criteria[12] . Chronic Kidney disease (CKD) was defined by persistent eGFR□< □60□ml/min/1.73□m^2^(CKD-EPI). The full hospital laboratory database was used to evaluate baseline creatinine values. AKI was only associated with the viral infection if the occurrence of AKI was within 72h after diagnosis of Rotavirus infection. Rotavirus infection was counted as nosocomial if diagnosed later than 72h after admission. Full remission (FR) of acute kidney failure was defined by serum creatinine values returning to baseline (but no worse than 20% above baseline). Partial remission (PR) was defined by serum creatinine values falling above 50% of maximal serum creatinine values, but have not met the FR criteria. The remaining cases were defined as no remission (NR).

### Microbiological rotavirus diagnostics

To detect rotavirus in stool samples, the CE-certified RIDASCREEN Rotavirus Antigen ELISA Test (R-Biopharm AG, Darmstadt, Germany) was performed. A stool sample of 50 - 100 mg was mixed with 1 ml of sample diluent for stabilization and liquefaction. The samples were then fully automated processed on a DSX-ELISA platform (DYNEX Technologies, Chantilly, USA) according to manufacturer’s instructions. Usually the samples were analyzed within 24h after arrival in the laboratory. For a few samples, there were delays in measurement for up to 24 h due to holidays or weekends.

### Study Outcomes

The primary outcome of the study was occurrence of acute kidney injury related to rotavirus infection. Secondary outcomes were incidence of renal failure in AKI stages I, II, and III, necessity of renal replacement therapy, in-hospital mortality and duration of hospitalization.

### Statistical Analysis

To describe the data, a descriptive statistic is first used: mean, standard deviation (SD), median, 1st quartile (Q1), 3rd quartile (Q3) as well as minimum (min), maximum (max) and sample size (N) are calculated for continuous variables (such as age, serum creatinine, eGFR, etc.); Numbers and percentages were calculated for categorical variables (e.g. gender, comorbidities, etc.). The descriptive statistics are collected for patients as a whole, as well as separately for patients with and without community-acquired infection and separately for patients with and without acute kidney injury (AKI). For the descriptive comparison of the survival times of patients with and without AKI, the Kaplan–Meier procedure was used. The log-rank test served as a significance test for comparison between the two studied groups, and the hazard ratio (HR) served as a descriptive measure of the difference in survival times. Data analysis of patient characteristics was performed using the chi-square test for categorical variables and t tests for continuous variables in cases where the data were normally distributed or the Wilcoxon test otherwise. Statistical analyses were performed using R statistical software (version 4.1.2) with implementation of the PS package (MatchIt). Any p-values that are less than 0.05 are considered statistically significant.

## Results

Overall, 314 hospitalized adult patients with rotavirus infection patients were evaluated, including 163 (51.9%) men and 151 women (48.1%). The mean age of the patients was 73.2 (±□15.6) years with 222 (70.7%) of patients older than 70 years. The dataset included 124□patients (39.5%) with diabetes mellitus, 105□patients (33.4%) with chronic kidney disease (CKD 3-5), 65 patients (20.7%) with heart failure, 81 patients (25.8%) with coronary heart disease, 28□patients (8.9%) with malignant disease and 13□patients (4.1%) with renal replacement therapy (RRT). Baseline patient characteristics and comorbidities are listed in **Table 1**.

**Table 1:** Patient characteristics of the rotavirus infection cohort, stratified according to the site of infection.

|  | Community-acquired | nosocomial | P value | overall |
| --- | --- | --- | --- | --- |
| **Number of patients** | 200 | 114 |  | 314 |
| **Age, years (SD)** | 71.2 (17.5) | 76.5 (10.8) | 0.004 | 73.1 (15.6) |
| **Age groups (%)** |  |  |  |  |
| <50 years n (%) | 23 (11.5) | 2 (1.8) | 0.013 | 25 (8.0) |
| 50-59 years n (%) | 15 (7.5) | 10 (8.8) |  | 25 (8.0) |
| 60-69 years n (%) | 29 (14.5) | 13 (11.4) |  | 42 (13.4) |
| >70 years n (%) | 133 (66.5) | 89 (78.1) |  | 222 (70.7) |
| **sex, male n (%)** | 97 (48.5) | 66 (57.9) | 0.138 | 163 (51.9) |
| **CKD n (%)** | 67 (33.5) | 38 (33.3) | 0.086 | 105 (33.4) |
| CKD stage 3 n (%) | 55 (27.5) | 31 (27.2) |  | 86 (27.4) |
| CKD stage 4 n (%) | 4 (2.0) | 6 (5.3) |  | 10 (3.2) |
| CKD stage 5 n (%) | 8 (4.0) | 1 (0.9) |  | 9 (2.9) |
| **Charlson Comorbidity Index** | 1.16 (1.4) | 1.68 (1.3) | 0.001 | 1.34 (1.4) |
| **Co-morbidities** |  |  |  |  |
| Diabetes mellitus n (%) | 79 (39.5) | 45 (39.5) | 1.000 | 124 (39.5) |
| Hypertension n (%) | 17 (8.5) | 5 (4.4) | 0.253 | 22 (7.0) |
| Heart failure n (%) | 27 (13.5) | 38 (33.3) | <0.001 | 65 (20.7) |
| Malignoma n (%) | 8 (4.0) | 20 (17.5) | <0.001 | 28 (8.9) |
| Coronary heart disease n (%) | 50 (25.0) | 31 (27.2) | 0.770 | 81 (25.8) |
| Peripheral artery disease n (%) | 12 (6.0) | 10 (8.8) | 0.487 | 22 (7.0) |
| Renal replacement therapy n (%) | 4 (2.0) | 9 (7.9) | 0.026 | 13 (4.1) |

### Community-acquired or nosocomial rotavirus infections

The majority of patients (200 patients, 63.7%) had community-acquired rotavirus infection, which was diagnosed at the time of hospital admission (Table 1). Our cohort also included 114 patients (36,3%) with nosocomial rotavirus infection. Patients with nosocomial infection were significantly older than patients with community-acquired infection (76.5 ± 10.8 years vs. 71.3 ± 17.5) years, p=0.004) and had a higher Charlson Comorbidity Index (1.68 ± 1.31 vs 1.16 ± 1.36, p=0.001). Overall, more patients with nosocomial rotavirus infections had heart failure (38 (33.3% vs 27 (13.5%), p<0.001) and malignant diseases (20 (17.5%) vs 8(4.0%), p<0.001).

### Acute kidney injury in rotavirus infections

Overall, 127 (40.4%) patients experienced acute kidney injury (AKI). Time-associated acute kidney injury occurred much more frequently in patients with community-acquired infection than in patients with nosocomial rotavirus infection (110 (55.0%) vs 17 (14.9%), p<0.001). 85.1% of 114 patients with nosocomial rotavirus infection did not experience acute kidney injury (AKI) (**Table 2**). Overall, 70 (55.1% of all patients with AKI) had AKI stage I, 22 (17.3%) patients had AKI stage II and 35 (27.6%) of patients experienced AKI stage III. The necessity of renal replacement therapy was significantly more frequent in the nosocomial infection cohort (9 (7.9%) vs 4 (2.0%) patients, p=0.026). The course of serum creatinine and urea during hospitalization in patients with community-acquired and nosocomial acquired infections is depicted in **Suppl. Figure S1**.

**Table 2:**
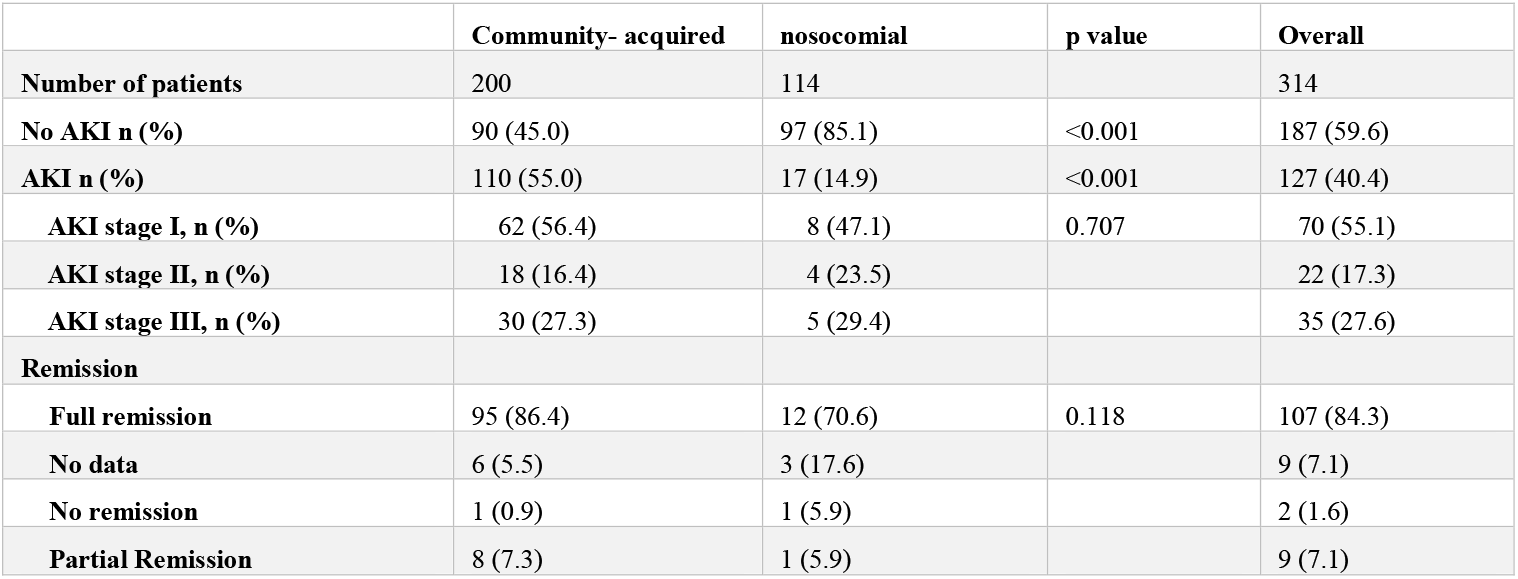
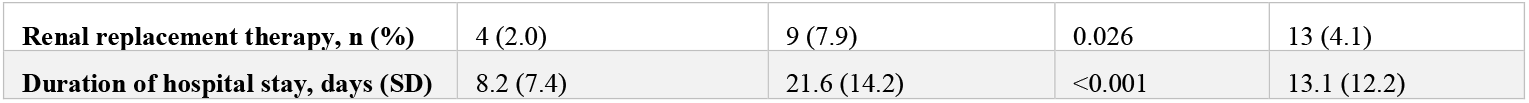
Occurrence of acute renal failure in the rotavirus infection cohort, stratified by the location of infection (community-acquired versus nosocomial).

**Suppl. Table S1** shows the baseline characteristics and comorbidities of patients with and without AKI. AKI (n=127) was most common in patients with pre-existing chronic kidney disease (AKI vs. no AKI, 46.5% vs. 24.6%; p<0.001). The majority of patients (n=107, 84.3%) with AKI in association with rotavirus infections were achieving renal remission **(Table 2)** with no significant differences between patients with community-acquired and nosocomial infection (95 (86.4%) vs. 12 (70.6), p=0.118).

### Length of hospitalization, in-hospital mortality and survival

Mean duration of hospital stay was 8.2 ± 7.4 days in community-acquired infection and 21.6 ± 14.2 days in patients with nosocomial infection (p<0.001). AKI did not significantly prolong the duration of hospital stay (11.6 ± 11.1 days in patients with AKI versus 14.0 ± 12.9 days in patients without rotavirus infection associated AKI, p=0.088). Overall, 26 (8.3%) of the rotavirus infection cohort died in hospital **(Table 3)**. In-hospital mortality (15 (11.8%) vs 11 (5.9%), p=0.096) was higher in the group with AKI versus without AKI. In addition, a survival analysis was performed on the full cohort (n = 314). The Kaplan–Meier survival curve revealed a significant difference between the two patient groups (see **Figure 1**). Patients with AKI experienced poorer survival outcomes (HR 2.63 (CI 1.20, 5.74) p=0.01).

**Table 3:** duration of hospitalization and in-hospital mortality.

|  | AKI | No AKI | p value | overall |
| --- | --- | --- | --- | --- |
| n | 127 | 187 |  | 314 |
| Duration of hospitalization, days | 11.6 (11.1) | 14.0 (12.9) | 0.088 | 13.1 (12.2) |
| In-hospital mortality n (%) | 15 (11.8) | 11 (5.9) | 0.096 | 26 (8.3) |

**Figure 1:**
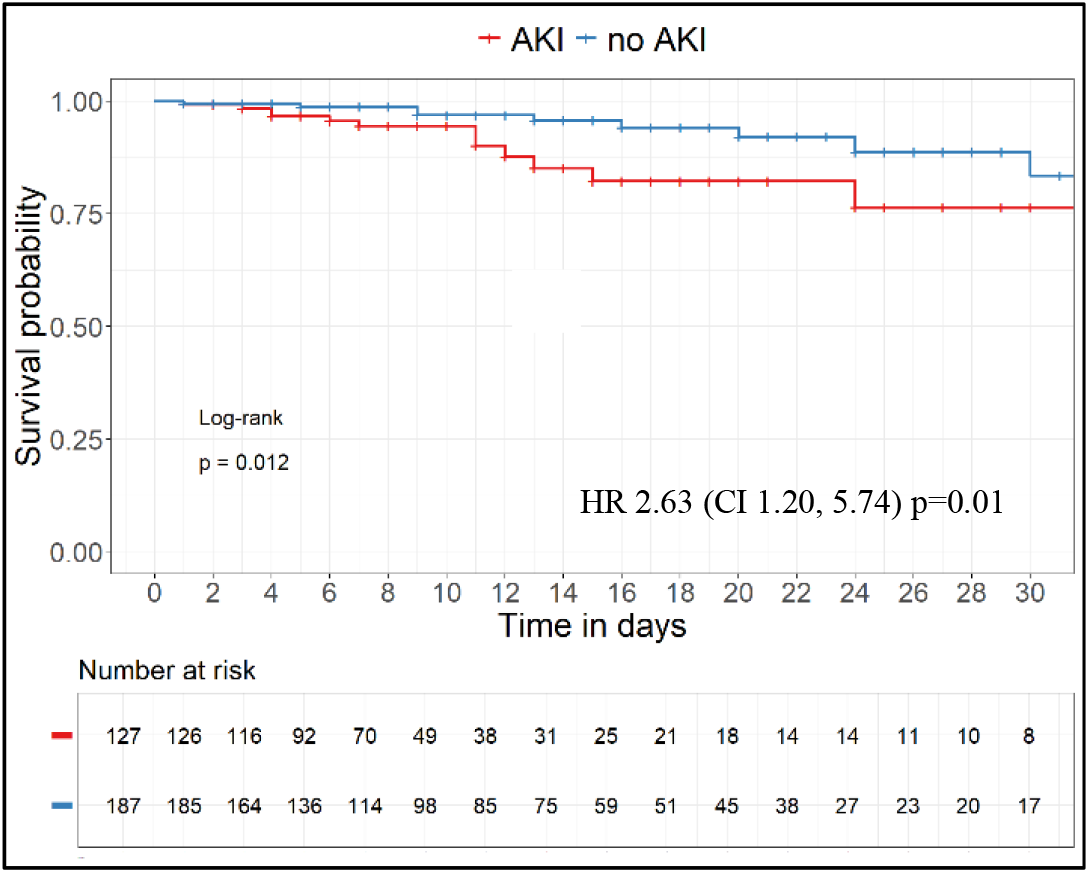
Kaplan-Meier survival curves with and without AKI.

### Sodium disorders and further laboratory parameters

Serum sodium levels were available in 65% (204/314) of patients on the day of rotavirus diagnosis (+/-1 day). 196 (96%) of these serum sodium levels were determined in patients with community-acquired rotavirus infections. In patients with community-acquired rotavirus infections, the mean serum sodium concentration was 136.8 ± 4.9 mmol/l. Hyponatremia was detected in 60 (30.6%) patients with a mean serum sodium concentration of 131.6 ± 3.6 mmol/l **(Table 4)**. 10 (5.1%) patients had serum sodium levels below 130 mmol/l with moderate and severe hyponatremia. Hyponatremia was significantly more common in patients with AKI (39.1% of 110 patients with AKI) compared to only 17 (19.8% of 86) patients without AKI, p=0.006.

**Table 4:**
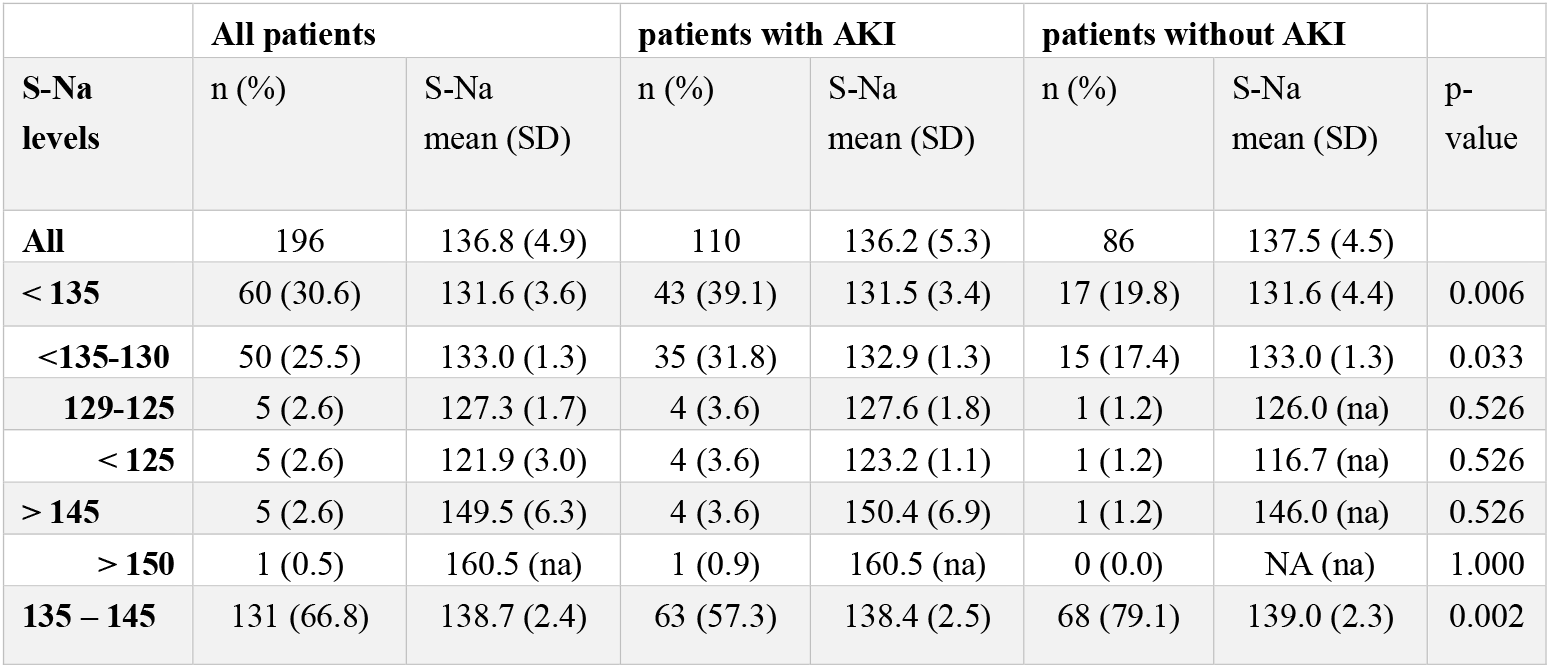
Sodium levels in 196 patients with community-acquired rotavirus infection. All serum sodium values are given as mean in mmol/l. SD: standard deviation. Serum sodium levels in mmol/l. p-values for numbers of patients with given serum sodium levels with AKI vs no AKI. na: not applicable. S-Na: serum-sodium. SD: standard deviation

|  | All patients |  | patients with AKI |  | patients without AKI |  |  |
| --- | --- | --- | --- | --- | --- | --- | --- |
| S-Na levels | n (%) | S-Na mean (SD) | n (%) | S-Na mean (SD) | n (%) | S-Na mean (SD) | p-value |
| All | 196 | 136.8 (4.9) | 110 | 136.2 (5.3) | 86 | 137.5 (4.5) |  |
| < 135 | 60 (30.6) | 131.6 (3.6) | 43 (39.1) | 131.5 (3.4) | 17 (19.8) | 131.6 (4.4) | 0.006 |
| <135-130 | 50 (25.5) | 133.0 (1.3) | 35 (31.8) | 132.9 (1.3) | 15 (17.4) | 133.0 (1.3) | 0.033 |
| 129-125 | 5 (2.6) | 127.3 (1.7) | 4 (3.6) | 127.6 (1.8) | 1 (1.2) | 126.0 (na) | 0.526 |
| < 125 | 5 (2.6) | 121.9 (3.0) | 4 (3.6) | 123.2 (1.1) | 1 (1.2) | 116.7 (na) | 0.526 |
| > 145 | 5 (2.6) | 149.5 (6.3) | 4 (3.6) | 150.4 (6.9) | 1 (1.2) | 146.0 (na) | 0.526 |
| > 150 | 1 (0.5) | 160.5 (na) | 1 (0.9) | 160.5 (na) | 0 (0.0) | NA (na) | 1.000 |
| 135 – 145 | 131 (66.8) | 138.7 (2.4) | 63 (57.3) | 138.4 (2.5) | 68 (79.1) | 139.0 (2.3) | 0.002 |

Hypernatremia (Dehydration) occurred in 5 (2.6%) patients (mean sodium concentration 149.5 ± 6.3mmol/l) with only one patient with serum sodium above 150mmol/l. 80% of all patients with hypernatremia were in the AKI group **(Table 4)**.

## Discussion

This is to the best of our knowledge the first report on incidence and outcome of acute renal injury (AKI) in adult hospitalized patients with rotavirus infections. There are only case reports on renal impairment in patients with rotavirus infections and exclusively in infants and children. In our cohort, acute renal deterioration was common with more than 40% of all patients experiencing AKI. The majority of AKI was graded AKI stage I (55.1%), but 27.6% of all patients have undergone AKI stage III and 4.1% even needed renal replacement therapy. Bogari et al analysed the incidence of AKI in a hospitalized cohort of patients with acute gastroenteritis in Saudi Arabia with only 13.6% of patients experiencing AKI[13] . Only 4.7% of this cohort had rotavirus infection. The most common pathogen was Salmonella, but AKI was most frequently reported in patients with Clostridioides difficile infection. This cohort is not comparable to the population of our study, not only because of the low frequency of rotavirus infections, but also due their inclusion of children (42.7% of the cohort) and the low representation of older adults (only 22.3% of patients were above the age of 60).

The higher vulnerability of the patients in our cohort is expressed in their old age and multiple comorbidities. More than 70% of all patients were above the age of 70 years and one-third of the cohort had pre-existing chronic kidney disease (CKD). Other frequent comorbidities included diabetes mellitus (39.5%), heart failure (20.7%) and coronary heart disease (25.8%).

Patients with AKI had a worse survival (HR 2.63 (CI 1.20, 5.74) p=0.01) compared to patients without AKI, which was not unexpected and most likely also due to a higher comorbidity burden. Regarding differences in baseline comorbidities, patients with AKI were more likely to have CKD compared to the patients with rotavirus infections without AKI. The majority of patients had community-acquired rotavirus infection and AKI was far more common in this cohort (55%) compared to patients with nosocomial infection (14.9%), which is most likely related to unresolved volume depletion. In hospitalized patients persisting volume depletion is less likely and treatment including volume expansion is usually undergone in a timely fashion. The majority of patients with AKI were achieving full remission after AKI, which hints to a mainly prerenal cause of AKI in most cases without significant ischemic components. Creatinine was significantly higher only until day 4 compared to patients without AKI in the community-acquired cohort (Figure 1). The course of creatinine in patients with nosocomial infection with AKI shows prolonged AKI which could be related to a greater ischemic component. Ischemic renal failure is an often-overlooked entity, frequently with a prerenal trigger, which leads to ischemia of the proximal tubule (and possible acute tubular necrosis) because of reduction in oxygen delivery through reduced blood supply in patients with ongoing volume depletion and blood pressure reduction. The reduction in blood flow in the peritubular capillaries, which are supplied by the post-glomerular postcapillary efferent arterioles, can be often aggravated by intake of RAS inhibitors or NSAIDs[14]. Patients with nosocomial rotavirus infection were significantly older and had significantly more often heart failure and malignancies as comorbidities, which might explain the higher need to initiate renal replacement therapy. Still, the frequency of achieving remission was not significantly different in patients with community-acquired versus nosocomial rotavirus infections.

The fact that most (84.3%) patients with AKI in association with rotavirus infections went into renal remission should not underestimate the significance of the kidney injury. AKI is a risk factor for the development of chronic kidney disease (CKD), even after apparent full remission. A systematic review and meta-analysis found that individuals with a history of AKI have an increased risk of developing new or progressive CKD[15] and this risk persists even when kidney function has returned to baseline levels. Another study demonstrated that the risk of renal progression remains elevated for up to ten years after an AKI episode, regardless of the severity of the initial injury[16] . This suggests that even if kidney function normalizes, the risk of CKD progression is not eliminated. Even though kidney disease progression is significantly more common in patients with a history of AKI compared to those without, this progression is mainly attributed to incomplete recovery of kidney function within the first months post-AKI[17]. These findings highlight that AKI serves as a significant risk factor for CKD, even if they achieve apparent full recovery of kidney function. Therefore, prevention of rotavirus infection associated AKI is of high importance.

Another interesting observation is the unexpectedly low incidence of dehydration in this vulnerable and rather geriatric cohort. Dehydration is caused by a significant water deficit and defined by the occurrence of hypernatremia. Hyponatremia was the most common electrolyte disorder in our cohort. Only very few cases of serum sodium above 145mmol/l were observed. Therefore, other than in children, dehydration is not a major concern in the adult population with rotavirus infection, even in comorbid old patients and dehydration is not linked to the high incidence of AKI. On the contrary, hyponatremia is common amongst adult comorbid patients with rotavirus infection. It can be assumed that hyponatremia is linked to volume depletion with (appropriate) increase in secretion of antidiuretic hormone (ADH). From this observation one can draw the conclusion that the most frequently made recommendation to increase the amount of water intake by these patients is certainly not helpful nor justified and may only increase and aggravate hyponatremia. Extracellular volume is not significantly influenced or maintained by water intake. The amount of sodium regulates extracellular volume and sodium intake is key to expansion of extracellular including intravascular volume in patients with rotavirus infection and volume depletion with high risk of developing AKI. Therefore, the recommendation should be to increase sodium intake (e.g. salty soups, salty dishes, etc.) instead of pure water intake in adult patients suspected of rotavirus infection und high risk of AKI.

## Limitations

Certain limitations of this study should be acknowledged. When concluding on potential management of patients with community acquired rotavirus infections, we only report data of patients being admitted to a hospital. The retrospective design with data from a single centre may influence the generalisability of findings. The causality of the association between rotavirus infection and AKI cannot be proven. We addressed this issue by only counting AKI when associated with rotavirus infection within 72h.

## Conclusion

Patients with rotavirus infections and certain risk factor (age > 70 years and comorbidities, e.g. CKD) have a high risk of developing AKI. Outpatients with typical gastrointestinal symptoms suggestive of rotavirus infection and risk factors should seek medical attention with a low threshold and, if necessary, undergo hospitalization to counteract volume depletion and the development of acute renal failure. Hyponatremia is common but dehydration rarely occurs in adult patients. Recommendations in outpatients at risk for AKI should focus on increasing salt intake for expansion of extracellular volume rather than water intake.

## Data Availability

All data produced in the present study are available upon reasonable request to the authors

## Author Contributions

UH, AR and RW were responsible for conceptualization. UH, AR, ON, KM and RW conceived the study and curated the data. KM cleaned, analyzed and visualized the data. All authors commented on the paper, oversaw the analysis, and edited the final manuscript. All authors contributed to important intellectual content and revision of the manuscript.

## Supplementary information

**Suppl. Table S1:**
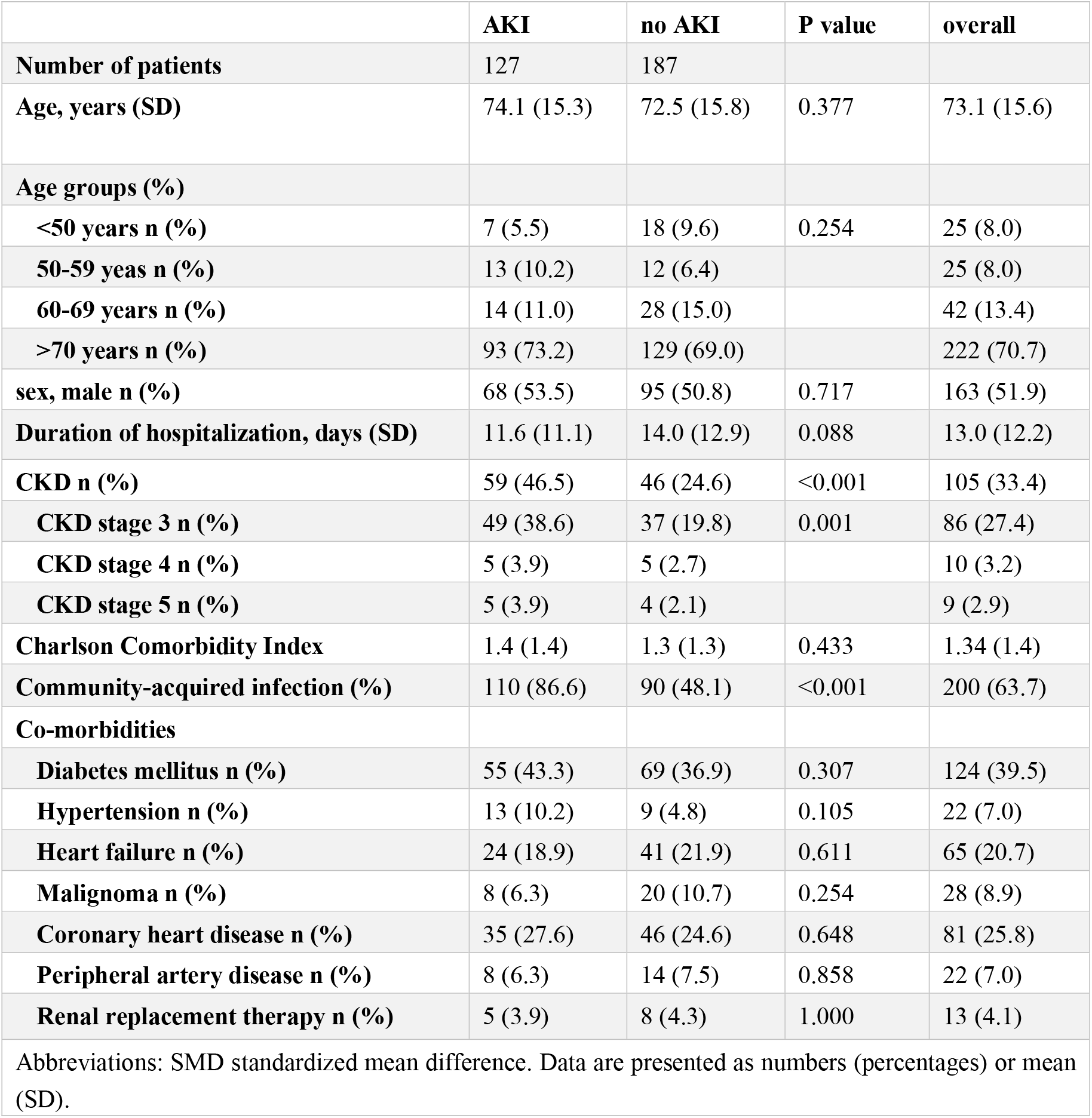
Baseline characteristics of patients with and without AKI.

**Suppl. Figure S1:**
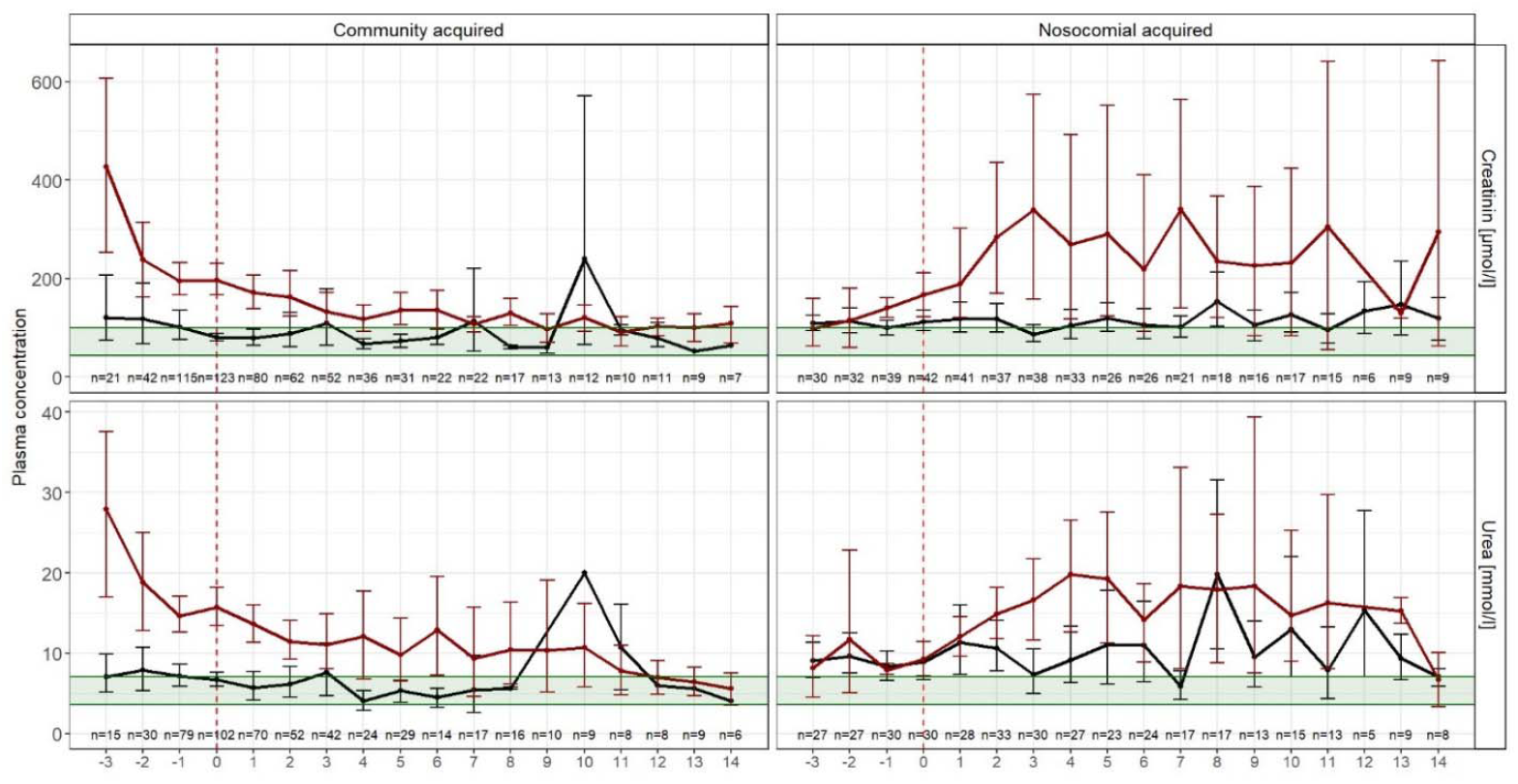
course of serum creatinine and urea in patients with community-acquired and nosocomial acquired infections 3 days before until max.18 days after rotavirus diagnosis. The day of rotavirus diagnosis is day 0. Means with 95% confidence intervals (black line: no AKI, red line = AKI, green area = normal range).

