## Supplementary material for "Acute kidney injury and hyponatremia in hospitalized patients with rotavirus infection": Suppl

**Supplementary information**

**Suppl. Table S1:** Baseline characteristics of patients with and without AKI

|  | AKI | no AKI | P value | overall |
| --- | --- | --- | --- | --- |
| Number of patients | 127 | 187 |  |  |
| Age, years (SD) | 74.1 (15.3) | 72.5 (15.8) | 0.377 | 73.1 (15.6) |
| Age groups (%) |  |  |  |  |
| <50 years n (%) | 7 (5.5) | 18 (9.6) | 0.254 | 25 (8.0) |
| 50-59 yeas n (%) | 13 (10.2) | 12 (6.4) |  | 25 (8.0) |
| 60-69 years n (%) | 14 (11.0) | 28 (15.0) |  | 42 (13.4) |
| >70 years n (%) | 93 (73.2) | 129 (69.0) |  | 222 (70.7) |
| sex, male n (%) | 68 (53.5) | 95 (50.8) | 0.717 | 163 (51.9) |
| Duration of hospitalization, days (SD) | 11.6 (11.1) | 14.0 (12.9) | 0.088 | 13.0 (12.2) |
| CKD n (%) | 59 (46.5) | 46 (24.6) | <0.001 | 105 (33.4) |
| CKD stage 3 n (%) | 49 (38.6) | 37 (19.8) | 0.001 | 86 (27.4) |
| CKD stage 4 n (%) | 5 (3.9) | 5 (2.7) |  | 10 (3.2) |
| CKD stage 5 n (%) | 5 (3.9) | 4 (2.1) |  | 9 (2.9) |
| Charlson Comorbidity Index | 1.4 (1.4) | 1.3 (1.3) | 0.433 | 1.34 (1.4) |
| Community-acquired infection (%) | 110 (86.6) | 90 (48.1) | <0.001 | 200 (63.7) |
| Co-morbidities |  |  |  |  |
| Diabetes mellitus n (%) | 55 (43.3) | 69 (36.9) | 0.307 | 124 (39.5) |
| Hypertension n (%) | 13 (10.2) | 9 (4.8) | 0.105 | 22 (7.0) |
| Heart failure n (%) | 24 (18.9) | 41 (21.9) | 0.611 | 65 (20.7) |
| Malignoma n (%) | 8 (6.3) | 20 (10.7) | 0.254 | 28 (8.9) |
| Coronary heart disease n (%) | 35 (27.6) | 46 (24.6) | 0.648 | 81 (25.8) |
| Peripheral artery disease n (%) | 8 (6.3) | 14 (7.5) | 0.858 | 22 (7.0) |
| Renal replacement therapy n (%) | 5 (3.9) | 8 (4.3) | 1.000 | 13 (4.1) |
| Abbreviations: SMD standardized mean difference. Data are presented as numbers (percentages) or mean (SD). | | | | |

**Suppl. Figure S1**: course of serum creatinine and urea in patients with community-acquired and nosocomial acquired infections 3 days before until max.18 days after rotavirus diagnosis. The day of rotavirus diagnosis is day 0. Means with 95% confidence intervals (black line: no AKI, red line = AKI, green area = normal range).

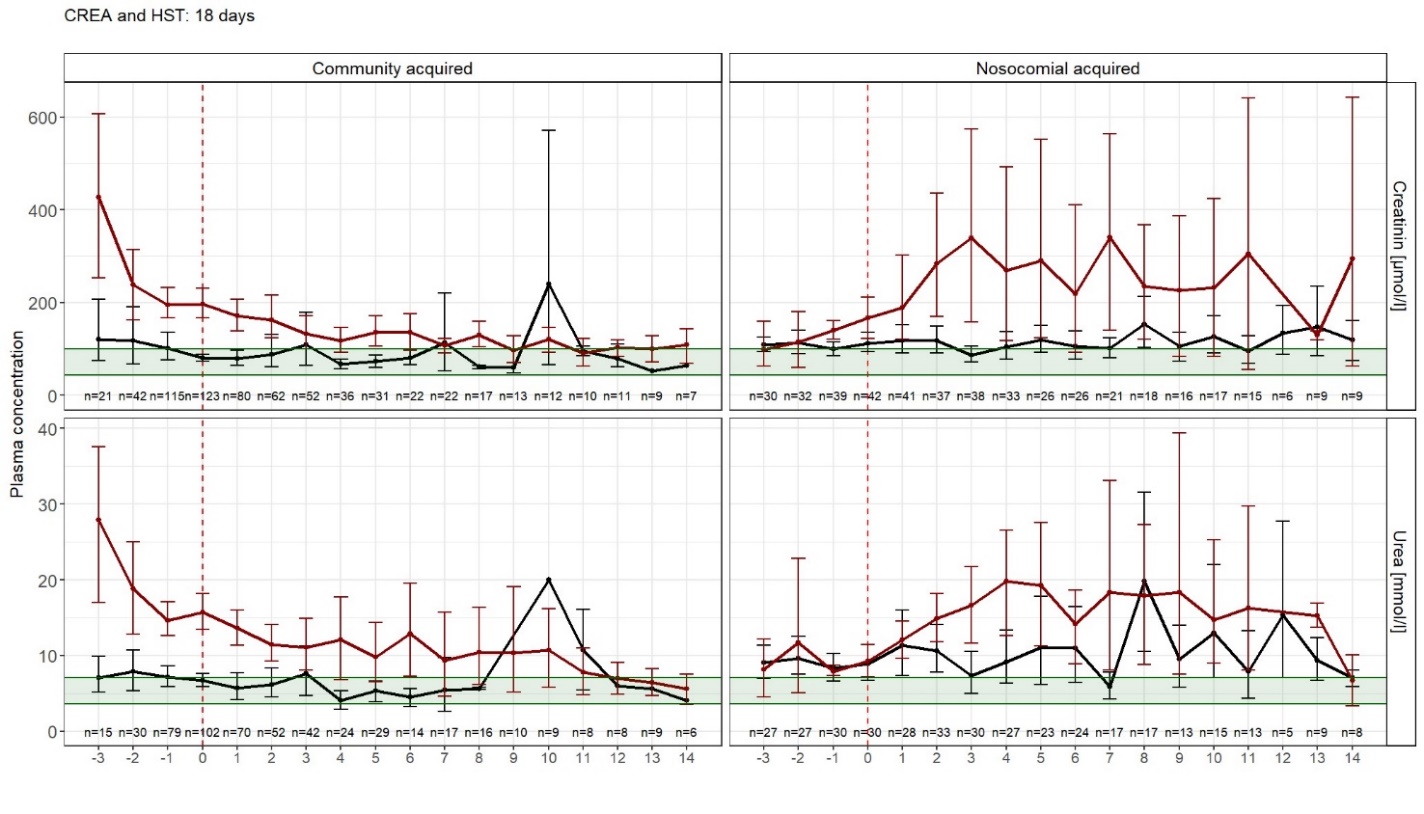
